# How optimal allocation of limited testing capacity changes epidemic dynamics

**DOI:** 10.1101/2020.12.21.20248431

**Authors:** Justin M. Calabrese, Jeffery Demers

## Abstract

Insufficient testing capacity continues to be a critical bottleneck in the worldwide fight against COVID-19. Optimizing the deployment of limited testing resources has therefore emerged as a keystone problem in pandemic response planning. Here, we use a modified SEIR model to optimize testing strategies under a constraint of limited testing capacity. We define pre-symptomatic, asymptomatic, and symptomatic infected classes, and assume that positively tested individuals are immediately moved into quarantine. We further define two types of testing. Clinical testing focuses only on the symptomatic class. Non-clinical testing detects pre- and asymptomatic individuals from the general population, and an “information” parameter governs the degree to which such testing can be focused on high infection risk individuals. We then solve for the optimal mix of clinical and non-clinical testing as a function of both testing capacity and the information parameter. We find that purely clinical testing is optimal at very low testing capacities, supporting early guidance to ration tests for the sickest patients. Additionally, we find that a mix of clinical and non-clinical testing becomes optimal as testing capacity increases. At high but empirically observed testing capacities, a mix of clinical testing and unfocused (information=0) non-clinical testing becomes optimal. We further highlight the advantages of early implementation of testing programs, and of combining optimized testing with contact reduction interventions such as lockdowns, social distancing, and masking.

## Introduction

The COVID-19 pandemic caught the world off-guard and continues to result in devastating consequences to life, health, and national economies. A key factor hampering global control efforts has been the unanticipated shortage of testing capacity. While testing was clearly problematic early in the pandemic, it remains a critical bottleneck in many parts of the world despite massive efforts to ramp up capacity (Hasell et al., 2020). Extensive testing provides the empirical basis on which to build a robust, scientifically based response strategy (Grassly et al., 2020). Insufficient testing leaves public health authorities with little information on how to coordinate efforts to effectively combat an emerging epidemic. For example, Li et al. (2020) estimated that, early in the COVID-19 outbreak in China, 86% of infections went undocumented, and these unnoticed cases fueled the subsequent global expansion of the disease. Similarly, undetected introductions of the virus coupled with undocumented community transmission facilitated the rapid spread of COVID-19 in New York City (Gonzalez-Reiche et al., 2020). Sustained high-rate testing also plays a crucial role in strategies for safely moving beyond costly and crippling lockdowns (Grassly et al., 2020). Specifically, quick identification and isolation of new infection clusters is critical for managing a disease like COVID-19 before a vaccine is widely available.

While other aspects of epidemic response such as vaccine distirbution have been studied from a resource allocation perspective (Zaric and Brandeau, 2001; Hansen and Day, 2011; Emanuel et al., 2020), the optimal allocation of limited testing capacity has, so far, received little attention (Grassly et al., 2020). The limited work that has been done on this topic has emerged recently, with some efforts focusing on using pooled testing as a simple means to stretch limited testing capacity as far as possible (Aragón-Caqueo et al., 2020; de Wolff et al., 2020; Ghosh et al., 2020; Gollier and Gossner, 2020; Jonnerby et al., 2020), while others consider stratified testing strategies focused on high risk groups such as health care workers (Cleevely et al., 2020; Grassly et al., 2020). Mathematical optimization has been applied to the economics of lockdown and quarantine policies (Aldila et al., 2020; Alvarez et al., 2020; Choi and Shim, 2021; Jones et al., 2020; Khatua et al., 2020; Piguillem and Shi, 2020), and to parameter estimation using testing data (Chatzimanolakis et al., 2020), but has not yet been applied comprehensively to resource allocation problems under testing constraints. Faced with insufficient testing capacity, public health agencies advise the prioritization of testing effort via qualitative considerations (Centers for Disease Control and Prevention, 2020). These guidelines base resource allocation decisions on total testing capacity, quality of information gained via contact tracing, current outbreak stage, and other characteristics specific to individual communities (Centers for Disease Control and Prevention, 2020). The proportion of limited testing resources reserved for high priority cases (e.g., highly symptomatic and vulnerable patients, essential healthcare workers) depends in part on the overall degree of sporadic versus clustered versus community-wide transmission (Robert Koch Institute, 2020; World Health Organization, 2020b). While these recommendations provide useful qualitative guidance, quantitative determination of optimal allocation strategies under limited testing is lacking despite its potential to increase testing efficiency.

Here, we address the optimal allocation of limited testing capacity with a COVID-19 specific SEIR ordinary differential equation compartmental model that features constrained testing and quarantine. We consider the allocation of testing and health care resources between two broad strategies (Centers for Disease Control and Prevention, 2020; Robert Koch Institute, 2020; World Health Organization, 2020b): 1) clinical testing focused on moderate to severely symptomatic cases, and 2) non-clinical testing designed to detect mildly symptomatic, pre-symptomatic, or fully asymptomatic cases. We further explore how the quality of proxy information with which to focus non-clinical testing on infected individuals affects the optimal balance between strategies. For both strategies, we assume that individuals that test positive are immediately moved into quarantine. We first quantify the extent to which an outbreak can be supressed via optimal testing and quarantine as a function of both testing capacity and proxy information quality. Specifically, we identify strategies that minimize the peak of the infection curve (i.e., “flatten the curve”). We then consider how positive factors like social distancing measures, and detrimental factors such as delays in testing onset affect optimal testing strategies and outbreak controllability. Throughout, we focus our analyses on empirically supported parameter values including realistic testing rates. While many existing COVID-19 SIR-like compartmental models explore the effects testing with forms of isolation like quarantine or hospitalization, the majority of these studies assume simple linear equations for the rates at which tests are administered and individuals are isolated (Adhikari et al., 2021; Ahmed et al., 2021; Amaku et al., 2021; Choi and Shim, 2021; Dwomoh et al., 2021; Hussain et al., 2021; Ngonghala et al., 2020; Rong et al., 2020; Saldanã et al., 2020; Sturniolo et al., 2021; Tuite et al., 2020; Verma et al., 2020; Youssef et al., 2021). We show (see Methods: Testing model) that linear models can not fully describe highly limited testing capacity scenarios, and we propose a novel, non-linear testing model that flexibly accounts for resource-rich and resource-limited settings.

## Methods

### Model development

We develop a modified SEIR model to determine how limits on the number of tests administered per day influence disease controllability, and to determine how limited resources can be best distributed among compartments in the modeled population. Our study was motivated by the COVID-19 crisis, both in terms of model structure, and in terms of the pressing need to make the most of limited testing capacity. Following other COVID-19 models (Contreras et al., 2020; Hellewell et al., 2020; Kretzschmar et al., 2020; Liu et al., 2020b; Piasecki et al., 2020; Rong et al., 2020), we assume two separate infectious categories based on observable symptoms. One, the “symptomatic class,” collects moderate to severely symptomatic cases for which one would typically seek clinical treatment, and the other, the “asymptomatic class,” collects all remaining cases, which may be either properly asymptomatic, or may simply be mild enough that the infected individual does not consider themselves sick or seek clinical treatment. We first develop a baseline disease model without interventions, and then incorporate testing and quarantine control strategies.

### Baseline SEIR model

We assume a homogeneously mixed population divided into *S* susceptible, *E* exposed, *A* asymptomatic and infectious, *Y* symptomatic and infectious, and *R* recovered classes. Newly infected individuals first enter the exposed class where they are unable to transmit the disease, and after a latent period, will enter the symptomatic or asymptomatic infectious class. For clarity, we take “asymptomatic” to include individuals who will show only mild to no symptoms over the course of the disease. The portion of individuals in the exposed class who eventually transition to the symptomatic class are considered “pre-symptomatic”. Although some evidence suggests that pre-symptomatic individuals can begin transmitting the disease one to several days before showing symptoms (Furukawa et al., 2020; He et al., 2020; Walsh et al., 2020), for simplicity, we assume that only *A* and *Y* class individuals are infectious. We further assume no host births or deaths, and that recovered hosts obtain permanent immunity. The model equations are as follows:

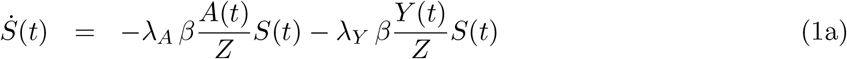

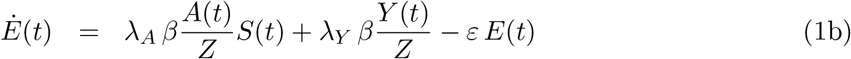

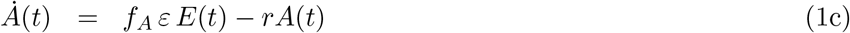

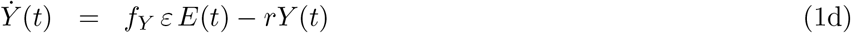

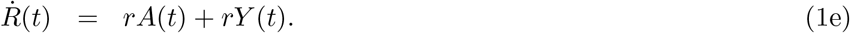

Here and throughout this paper, over dots denote derivatives with respect to time, and we measure time in units of days. The meaning of each model parameter, and the numerical values used, are given in Table 1. We note that while the recovery time 1/*r* and incubation period 1/*ε* can be consistently estimated from data, some parameters in our model not accurately known. Specifically, the fractions of asymptomatic and symptomatic infectious populations, *f*_*A*_ and *f*_*Y*_, respectively, are highly uncertain parameters, as estimates based on both modeling and clinical data place the truly asymptomatic population anywhere from 1% to 80% of all infections (Furukawa et al., 2020; Walsh et al., 2020; Widders et al., 2020). Focusing on symptomatic individuals, the fractions that are mildly symptomatic versus moderately to severely symptomatic are also uncertain, although some evidence suggests the majority of cases are mild (Liu et al., 2020a). Based on these observations, we choose *f*_*A*_ = 0.75 and *f*_*Y*_ = 0.25 as baseline values. Studies quantifying viral loads via RT-PCR tests and viral culture studies generally show that asymptomatic individuals are as, or less, infectious than symptomatic individuals (Walsh et al., 2020; Widders et al., 2020), and that more severely symptomatic cases can be associated with higher viral loads (Liu et al., 2020a; Walsh et al., 2020; Widders et al., 2020). We therefore assume that the symptomatic transmission probability, *λ*_*Y*_, is twice that of the asymptomatic transmission probability, *λ*_*A*_. Finally, we choose the overall values of *λ*_*A*_, *λ*_*Y*_, and the contact rate, *β*, to yield a basic reproduction number of *R*_0_ = 5.0 absent of any testing or quarantine control (see the Appendix for an analytic expression for *R*_0_). This *R*_0_ value falls roughly in the middle of the ranges suggested by a number of studies (Jiang et al., 2020; Majumder and Mandl, 2020; Rong et al., 2020; Sanche et al., 2020). Note that under our model parameters, in the absence of testing and quarantine, the symptomatic and asymptomatic contributions to *R*_0_ are 2.0 and 3.0, respectively.

**Table 1:** Model parameter definitions and baseline numerical values used. Values for highly uncertain parameters based on the current literature for which we make an estimate are indicated with an asterisk.

| Parameter | Name | Meaning | Value | Refs |
| --- | --- | --- | --- | --- |
| $\beta$ | Contact rate | Average number of contacts per individual per unit time | 4.0* | Jiang et al. (2020); Majumder and Mandl (2020); Rong et al. (2020); Sanche et al. (2020) |
| $\lambda_A$ | Asymptomatic transmission probability | Probability of disease transmission per susceptible-symptomatic contact | 0.125* | Jiang et al. (2020); Majumder and Mandl (2020); Rong et al. (2020); Sanche et al. (2020) |
| $\lambda_Y$ | Symptomatic transmission probability | Probability of disease transmission per susceptible-asymptomatic contact | $2\lambda_A^*$ | Liu et al. (2020a); Walsh et al. (2020); Widders et al. (2020) |
| $1/\varepsilon$ | Latent period or incubation period | Time between transmission and onset of infectiousness or symptoms | 5 days | Furukawa et al. (2020); Hellewell et al. (2020); He et al. (2020); Lauer et al. (2020); Rong et al. (2020); Sanche et al. (2020) |
| $f_A$ | Asymptomatic fraction | Fraction of infections which remain mild or asymptomatic | 0.75* | Furukawa et al. (2020); Grassly et al. (2020); Liu et al. (2020a); Walsh et al. (2020); Widders et al. (2020) |
| $f_Y$ | Severely symptomatic fraction | Fraction of infections which become severe and symptomatic | $1 - f_A^*$ | - |
| $1/r$ | Infectious period | Average time over which infected individuals can actively transmit the virus | 8 days | Walsh et al. (2020); Widders et al. (2020) |
| $Z$ | Population size | Total number of hosts (assumed fixed) | 50000 | Assumed |

### Testing model

To analyze testing and quarantine control strategies operating with testing capacity constraints, we construct a simple model that scales smoothly between extremes of abundant and severely constrained testing resources. This model is governed by the testing capacity, *C*, and the testing time, *τ*. The testing capacity *C* denotes the maximum achievable per capita testing rate assuming a fixed level of resources. This maximum testing rate represents the limitations of a finite health care infrastructure and finite testing supplies, and we take “increased resources” to mean an increased value of *C*. The testing time represents the average amount of time required for an individual be tested and obtain results, absent any backlogs or waiting times due to other patients. Time-consuming factors independent of the number of people needing to be tested determine the value of *τ*, for example, procrastination, travel time, and test processing times. Suppose that some sub-population *P* (*t*) of the total population *Z* is eligible to be tested at time *t*, and let 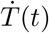 denote the rate at which tests are administered and processed for results. We propose the following expression:

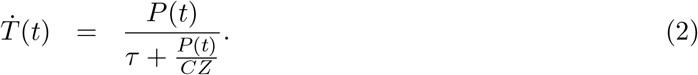

Under this model,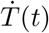 falls between two limiting expressions representing “resource-limited” and “testing time-limited” testing regimes, defined by 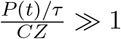 and ≪1, respectively:

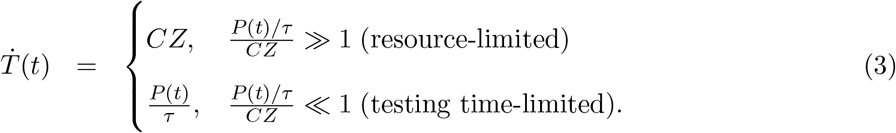

The testing time-limited regime represents a high resource availability scenario, where the total testing rate is limited only by the rate at which individuals arrive to be tested and the actual time required for a single test to produce results. The resource-limited regime represents a low resource availability scenario, where the number of people needing to be tested far exceeds the maximum daily testing capacity.

It is important to note that despite its frequent use in the literature, a simple linear testing rate model 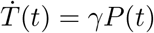, where *γ* represents a testing rate parameter, is insufficient for describing resource-limited scenarios. Under a linear model, even if *γ* is made to be very small in reflection of testing limitations, the rate at which tests are administered will always increase in proportion with the demand for testing, and this can not describe a resource-limited scenario where the maximum testing rate is capped at a fixed value independent of the testing demand. The linear model is in fact equivalent to our testing model in Eq. (2) under the testing time-limited regime shown in Eq. (3), which represents a resource-rich rather than resource-limited scenario.

We note further that the testing rate model in Eq. (2) can be extended to multiple sub-populations subject to distinct testing capacity constraints. Specifically, suppose two distinct sub-populations *P*_1_(*t*) and *P*_2_(*t*) are subject to two distinct testing policies with distinct resource pools limited by the capacities *C*_1_ and *C*_2_, respectively. In this scenario, the total rate at which tests are administered to the two populations is given by the following:

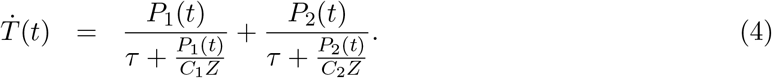

### Disease model with resource allocation, contact tracing, and quarantine

We now utilize our testing model to incorporate a quarantine and contact tracing program into our disease model. We assume that testing resources can be allocated between two control strategies, designated “clinical testing” and “non-clinical testing,” which are applied to individuals based the presence of observable symptoms. Clinical testing applies resources to the moderate to severely symptomatic class *Y* (*t*). This strategy represents saving resources for hospital and health care facilities to ensure adequate treatment of the most seriously ill individuals. Under a pure clinical testing strategy, individuals are only eligible to be tested if they show severe enough symptoms. Non-clinical testing applies resources to the exposed class *E*(*t*) and the asymptomatic class *A*(*t*), as well as to some portion of the uninfected population. This strategy represents a combination of contact tracing and population monitoring designed to identify asymptomatic individuals before they can transmit the disease. For both strategies, we assume perfectly accurate tests with no false positives or negatives, and we assume that testing can detect the disease at any point after infection to when the period of infectiousness ends. These assumptions are somewhat optimistic in comparison to real-world testing efficacies (Kucirka et al., 2020; Surkova et al., 2020), and our model thus represents a limit on what can be achieved.

When an infected individual is tested in our model, they will instantly transition to the quarantine class *Q* (*t*), and will subsequently recover from the disease and transition to the recovered class. We also introduce the “unknown status” class *U* (*t*), which is the subset of recovered hosts who did not receive any testing or quarantine, and are therefore unaware that they previously had COVID-19. We assume that the contact tracing and monitoring programs modeled by non-clinical testing cover the entirety of the *E*(*t*) and *A*(*t*) classes, and some fraction (1 − *η*) of the *S*(*t*) + *U* (*t*) class, where *η* ∈ [0, 1] denotes the “information parameter.” The value of *η* represents the quality of information gained through contact tracing and case investigations, where larger *η* represents better information, and the quantity (1 − *η*)(*S*(*t*) + *U* (*t*)) gives the fraction of the COVID-19 negative population with an unknown infection history which will be targeted under non-clinical testing. In other words, with better information, more non-clinical testing resources will be used for quarantining the COVID-19 positive population, and less will be “wasted” obtaining negative results. The case *η* = 1 represents perfect contact tracing and case investigation, where all non-clinical testing is focused on infected individuals. While this extreme is unrealistic, *η* values close to 1 may be plausible given high quality contact information and well-understood disease dynamics. The case *η* = 0 represents no information such that non-clinical testing is randomly dispersed among the non-symptomatic infected classes and the entirety of the *S*(*t*) + *U* (*t*) classes, and thus represents a large scale population monitoring program.

Suppose that a fraction *ρ* of the testing capacity *C* is allocated to non-clinical testing, with the remainder devoted to clinical testing. The parameter *ρ* denotes the “strategy parameter,” and its value represents a government’s policy for balancing health care resources between reservation for more critical symptomatic cases and for use in contact tracing or monitoring programs. Our modified SEIR model including testing, contact tracing, quarantine, and resource allocation is as follows:

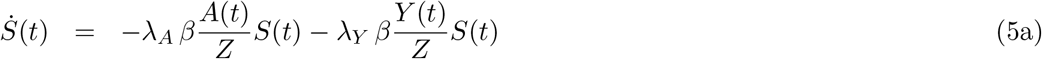

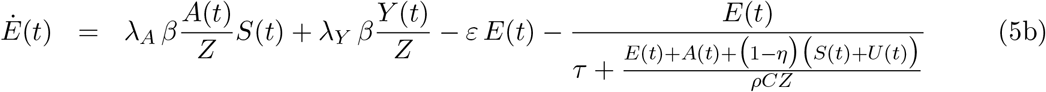

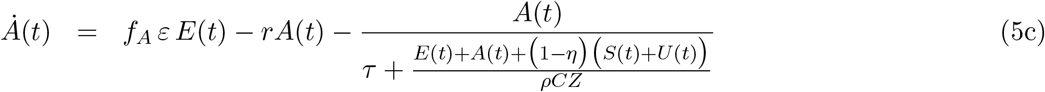

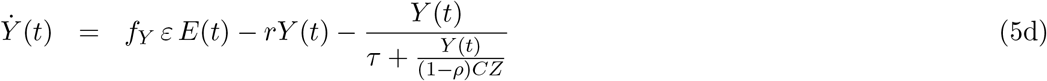

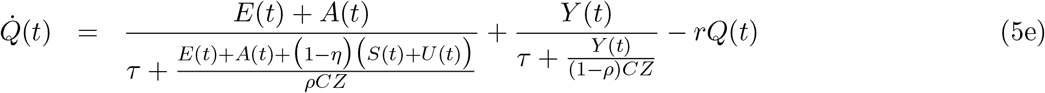

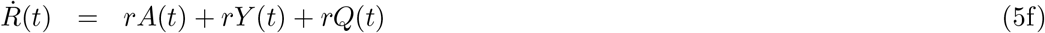

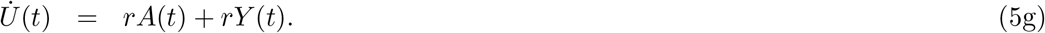

Note that as *ρ* → 1, Eq. (5d) reduces to Eq. (1d), and as *ρ* → 0, Eqs. (5b) and (5c) reduce to Eqs. (1b) and (1c), respectively. Additionaly, as *C* → 0, all of Eq. (5) reduces to Eq. (1). In the Appendix, we analyze a closed-form expression for *R*_0_ under our full SEIR + testing and quarantine model.

A summary of all control related parameters is given in Table 2 for reference. For all simulations, we assume the testing time *τ* = 1 day, which is reasonable for an effective testing and processing system lacking patient backlogs. For all other control parameters, we will consider a range of numerical values. Note that for notational simplicity in our model equations, we define *C* in units of tests per person per day, while actual testing capacities are often reported in units of tests per thousand people per day. To establish clear connections between our results and real-world testing limitations, we too report numerical values for testing capacity in units of tests per thousand per day. Thus, if we report a particular numerical value *C*_1*K*_ in per thousand units, the corresponding value in per person units used for numerical simulations is given by *C*_1*K*_/1000.

**Table 2:**
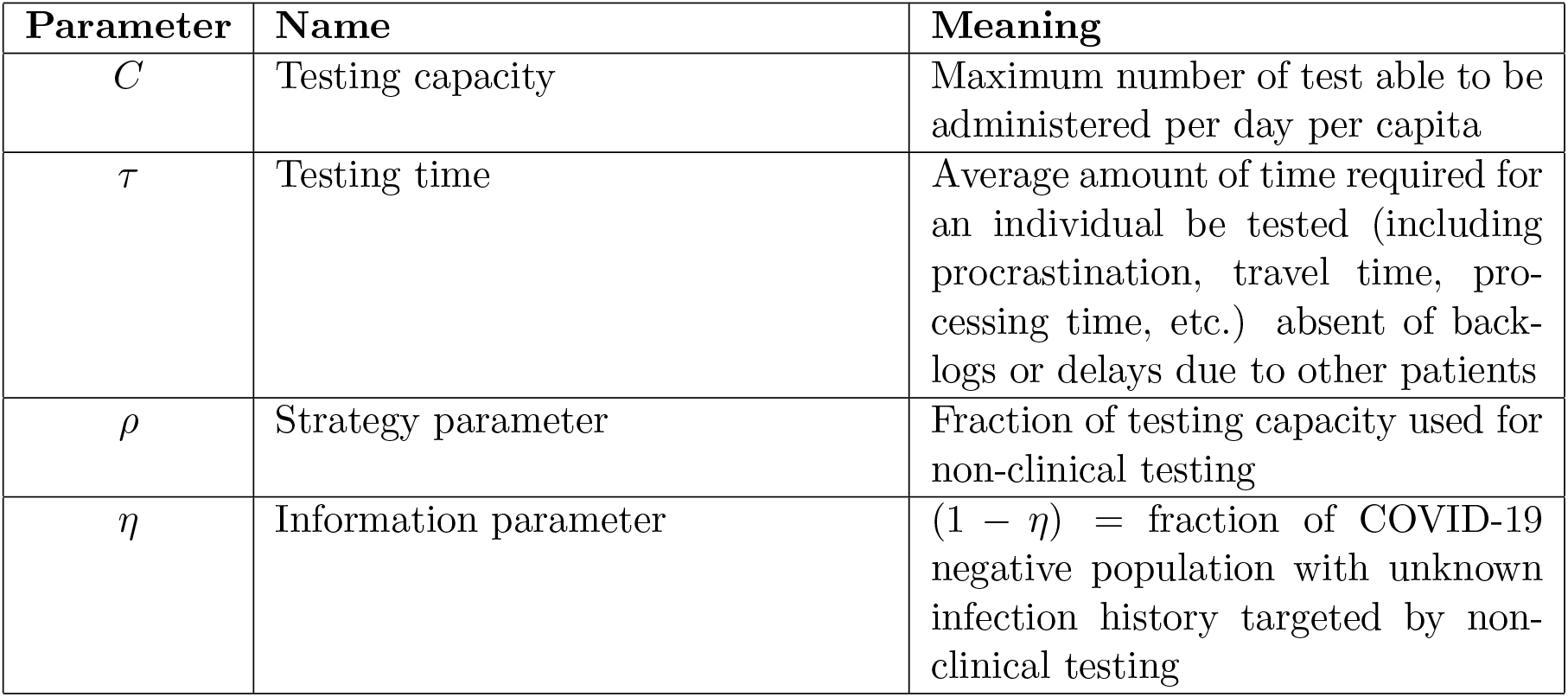
Testing and quarantine control parameter definitions

| Parameter | Name | Meaning |
| --- | --- | --- |
| $C$ | Testing capacity | Maximum number of test able to be administered per day per capita |
| $\tau$ | Testing time | Average amount of time required for an individual be tested (including procrastination, travel time, processing time, etc.) absent of backlogs or delays due to other patients |
| $\rho$ | Strategy parameter | Fraction of testing capacity used for non-clinical testing |
| $\eta$ | Information parameter | $(1 - \eta)$ = fraction of COVID-19 negative population with unknown infection history targeted by non-clinical testing |

### Flattening the epidemic peak as a control goal

In accordance with the goal of “flattening the curve” typically communicated by government and health agencies (World Health Organization, 2020a), we simulate our model dynamics to determine if and to what extent appropriately allocated resources can reduce the peak number of infections. First, we calculate optimal resource allocation strategies *ρ* for reducing the epidemic peak (defined as the maximum value of the sum of the *E, A*, and *Y* classes), assuming parameter values in Table 1 and an initial outbreak of one exposed individual as our baseline case. Optimization is executed by numerically integrating the disease dynamics in Eq. (5) and utilizing the *fmincon* function in *MatlabR2017a* running the *sqp* algorithm with *ρ* = 0 as an initial guess. To account for the possibility of multiple local minima, we employ the parallel *MultiStart* algorithm from *Matlab’s* global optimization toolbox. Simulations assume specific values values for *η* and find optimal *ρ* and epidemic peak values for all testing capacities in the range [0, 25] (in units of tests per thousand per day). In the Appendix, we consider the alternative optimization goal of minimizing *R*_0_ under our combined disease + testing model.

To determine the effects of delays in testing/quarantine policy implementation, as well as the effects of social distancing efforts, we consider alternative scenarios of initial conditions and/or model parameters. We model implementation delays by considering initial conditions equal to the outbreak size after a given number days under our baseline scenario with no testing or quarantine controls. In the case of a 30 day delay, for example, the alternative initial condition is given by (*S*(0), *E*(0), *A*(0), *Y* (0), *R*(0)) = (49727, 134, 63, 21, 55), which yields 218 initially infected individuals. To model the effects of social distancing, we reduce *β* to a given fraction of its baseline value. Additionally, we consider the effects of social distancing and implementation delays together. To evaluate the effects of alternative scenarios on optimal control policies, we perform the same optimization procedure as in our baseline case. We provide an in-depth examination of the specific conditions of a 30 day initial testing delay and a 50% reduction of *β*, and also consider a broader range of delays and *β* reductions in less detail.

## Results

### Optimal resource allocation strategies

We find that, even under extremely limited testing capacities, the epidemic peak can be reduced to the initial outbreak size of 1 infected individual, provided that resources are optimally allocated and sufficient information quality is available (Fig. 1a). Reducing the epidemic peak to the initial outbreak size signifies that disease spread has been effectively suppressed. For a given *η* at low testing capacities, the optimal strategy is to devote all resources to clinical testing, and a minimum threshold capacity *C*^*th*^(*η*) exists, above which optimal strategies call for a mix of clinical and non-clinical testing (Fig. 1b). As testing capacity increases above *C*^*th*^(*η*) optimal strategies require an increasing share of resources to be devoted to non-clinical testing until a second threshold capacity *C*\*(*η*) is reached. The threshold *C*\*(*η*) represents the smallest testing capacity for which the outbreak can be suppressed to its initial size with an information level *η*. For example, at information level *η* = 0.90, *C*^*th*^(*η*) = 2.8 and *C*\*(*η*) = 15.4 tests per thousand per day (Fig. 1b). Table 3 summarizes the threshold capacity definitions and gives numerical values for various values of *η*.

**Table 3:** Threshold testing capacity definitions and numerical values for the information levels *η* considered in Fig. 1. Note that critical information threshold levels *η*^*crit*^(*C*) can be inferred from this table from the relationship *C*^*th*^(*η*_0_) = *C*_0_ if and only if *η*^*crit*^(*C*_0_) = *η*_0_. For example, the *η* = 0.90 column indicates that *η*^*crit*^(2.8) = 0.90.

| Threshold Capacity | Definition | Numerical Values (tests per thousand per day) |  |  |  |  |  |  |  |
| --- | --- | --- | --- | --- | --- | --- | --- | --- | --- |
| | | $\eta = 0.00$ | $\eta = 0.50$ | $\eta = 0.85$ | $\eta = 0.90$ | $\eta = 0.95$ | $\eta = 0.97$ | $\eta = 0.999$ | $\eta = 1.00$ |
| $C^{th}(\eta)$ | Minimal capacity beyond which optimal strategies are mixed | 8.0 | 6.0 | 3.4 | 2.8 | 1.8 | 1.2 | 0.1 | 0.0 |
| $C^*(\eta)$ | Minimal capacity beyond which optimal strategies reduce epidemic peaks to initial infection levels | 154.0 | 77.0 | 23.1 | 15.4 | 7.6 | 4.6 | 0.2 | 0.0 |

**Figure 1:**
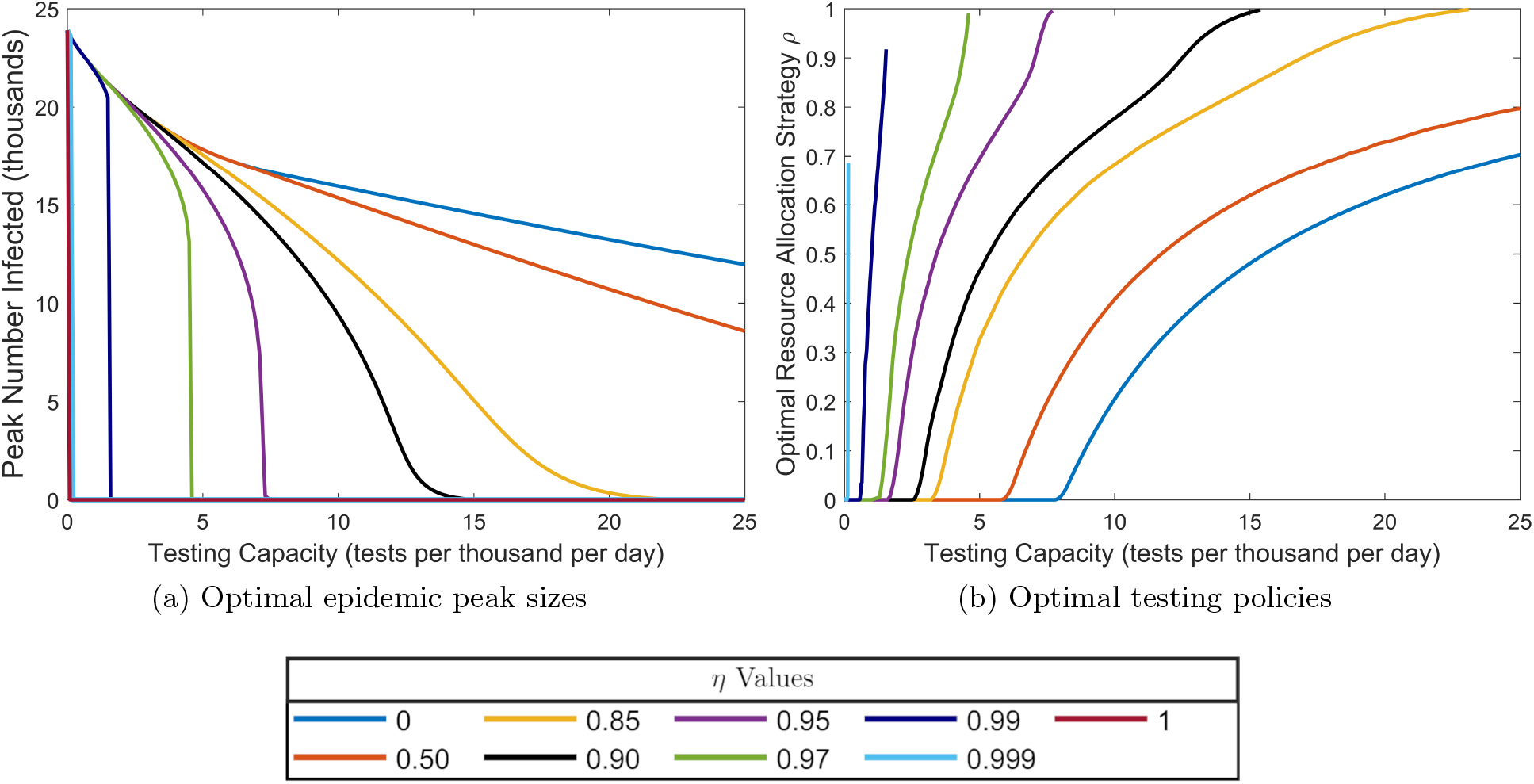
Optimally reduced epidemic peak sizes and corresponding optimal *ρ* values as a function of testing capacity. Note that the peak value 23,882 at zero testing capacity corresponds to the uncontrolled disease dynamics without a testing or quarantine program. **(a)** Optimally reduced epidemic peak sizes as a function of testing capacity for the values of non-symptomatic testing information quality *η* indicated in the legend. **(b)** Optimal resource allocation strategies *ρ* for reducing the epidemic peak as a function of testing capacity. An optimal *ρ* curve which terminates at a testing capacity *C*\* below than the maximally considered value 25.0 tests per thousand per day indicates an information level for which the optimal strategy is not unique at capacities above *C*\*, and for which the optimal epidemic peak size can be reduced to the initial value of one infected at capacities above *C*\*. Note that for perfect information *η* = 1, the optimal testing strategy is not unique down to the smallest non-zero testing capacity considered 0.01 tests per thousand per day. Note also that for *η* = 0.85, 0.90, 0.95, and 0.97, the optimal *ρ* values at *C* = *C*\* appear to be close to 1, but are not actually equal to 1.

For testing capacities *C > C*\*(*η*), the epidemic peak size will always be reduced to 1 as long as at least as much of the total capacity is devoted to clinical and non-clinical testing as is called for by the optimal strategy at *C* = *C*\*(*η*). As a result, optimal strategies are not unique when *C > C*\*(*η*). To see this non-uniqueness explicitly, consider an information level *η*, and let *ρ*\*(*η*) denote the optimal strategy parameter at the critical capacity *C*\*(*η*). At this critical capacity, the optimal action is to devote *ρ*\*(*η*)*C*\*(*η*) total resources to non-clinical testing and (1 − *ρ*\*(*η*)) *C*\*(*η*) total resources to clinical testing, the result of which reduces the epidemic to the smallest possible value 1. If the testing capacity *C* exceeds the critical level *C*\*(*η*), one can always allocate at least *ρ*\*(*η*)*C*\*(*η*) and (1 − *ρ*\*(*η*)) *C*\*(*η*) total resources to non-clinical and clinical testing, respectively, thereby guaranteeing the epidemic peak to be reduced to 1. The allocation of the remaining *C* − *C*\*(*η*) resources will therefore be irrelevant, as adding resources to either strategy can not further decrease the peak size beyond the initial infection size. In other words, for a given *η*, if *C > C*\*(*η*), the epidemic peak will be reduced to 1 whenever *ρ* is selected such that *ρC* ≥ *ρ*\*(*η*)*C*\*(*η*) and (1 − *ρ) C* ≥ (1 − *ρ*\*(*η*)) *C*\*(*η*). These inequalities imply that any *ρ* drawn from the interval 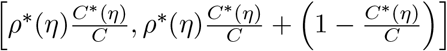 will reduce that epidemic peak to the minimum possible value, thus showing that the optimal strategy is not unique for *C > C*\*(*η*).

For a given capacity *C*, there exists a critical information value *η*^*crit*^(*C*), below which the optimal strategy is clinical testing only, and above which the optimal strategy is mixed (Fig. 1). From the definition of *C*^*th*^(*η*) as the minimal capacity below which the optimal strategy is clinical testing only for a given *η*, we have the relation *C*^*th*^(*η*_0_) = *C*_0_ if and only if *η*^*crit*^(*C*_0_) = *η*_0_, and numerical values for *η*^*crit*^(*C*) at specific *C* values can therefore be inferred from Table 3. The critical information value represents an important practical decision making threshold for public health officials operating under a potentially limited testing capacity *C*; if *η* is estimated to be below *η*^*crit*^(*C*), no testing resources should be diverted away from severely symptomatic individuals, while if *η* is estimated to be above *η*^*crit*^(*C*), important resource management decisions should be considered. In Fig. 2, we plot *η*^*crit*^ as a function of testing capacity *C*. Here, the curve defined by *η*^*crit*^(*C*) divides the (*C, η*) plane into two regimes, one where the optimal strategy calls for clinical testing only, one where optimal strategies are a mix of clinical and non-clinical testing. In particular, we find that for *C >* 8.0 tests per thousand per day, *η*^*crit*^(*C*) = 0. Thus, for testing capacities above 8.0, it is always optimal to devote at least some resources to non-clinical testing, even if the non-clinical testing is a simple randomized population sampling program lacking the efficacy of targeted contact tracing efforts.

**Figure 2:**
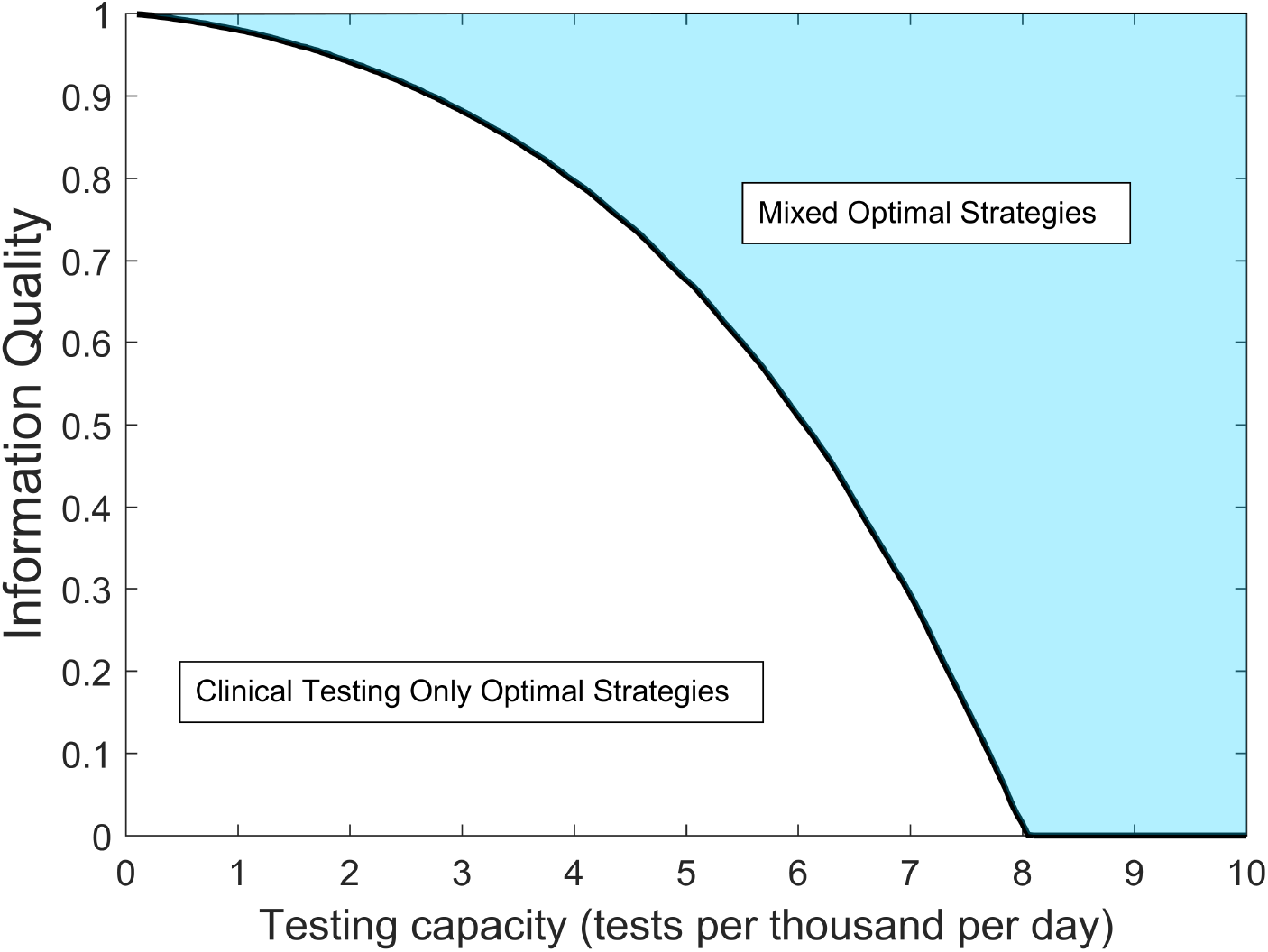
Optimal resource allocation strategy regimes for reducing the epidemic peak as a function of testing capacity *C* and non-clinical testing information quality *η*. For (*C, η*) values within the shaded region, optimal strategies call for sharing resources between a mix of clinical and non-clinical testing. Within the non-shaded region, optimal strategies call for all resources to be concentrated to clinical testing only. The black curve indicates a critical information threshold which for a given testing capacity, determines whether the optimal strategy will be mixed or clinical testing only.

### Social distancing and delays in testing program implementation

Unsurprisingly, delaying the implementation of a testing program by 30 days has negative impact on optimal peak reduction, with the delay being most detrimental at the lowest testing capacities (cf. Figs 3a and 3b). Specifically, a delay of this magnitude makes it impossible to reduce the epidemic peak to its initial value, regardless of the information level, within the range of testing capacities [0, 1.2] tests per thousand per day (Fig. 3a). This is not the case for immediate testing program implementation, where the peak can be reduced to its initial value at *any* non-zero testing capacity given sufficient information quality (Fig. 1). Reducing the peak to its initial value is an important control goal, as it is equivalent to the ability to force an immediate downturn in the infection curve upon implementation of testing and quarantine. These results emphasize the need for early implementation of a testing program at the beginning stages of a novel disease epidemic, where resources may be extremely limited as health agencies adjust to the biology of the newly discovered infectious agent.

**Figure 3:**
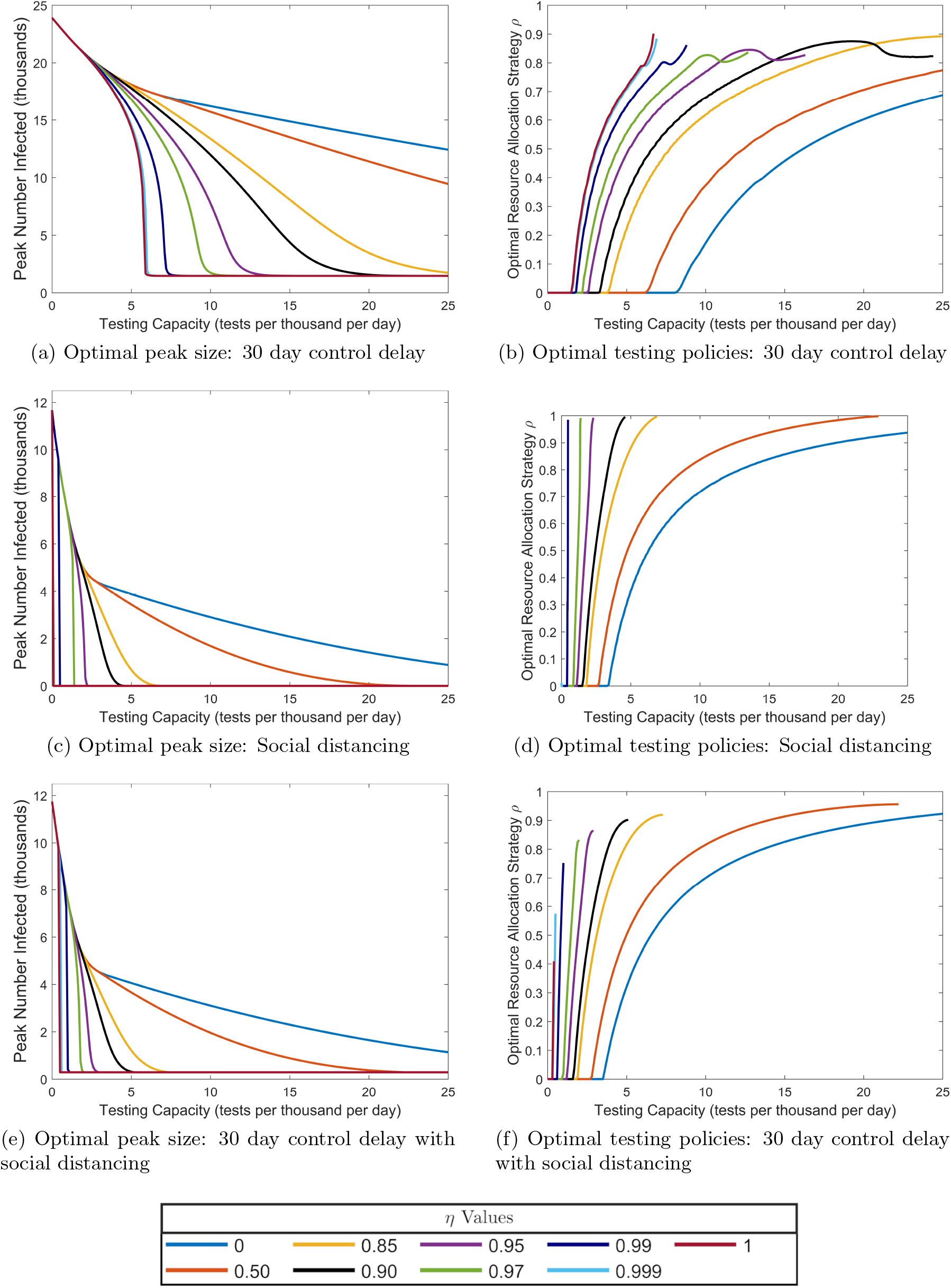
Effects of social distancing and control delays on optimal testing strategies for reducing the epidemic peak. See Fig. 1 for a comparison to our baseline case and an explanation of the meaning of each plot.

Halving the contact rate, which simulates the influence of social distancing, has a strong effect on optimal policies and peak sizes (Figs. 3c and 3d). At zero testing capacity (which corresponds to the disease dynamics without testing and quarantine), the epidemic peak reaches 11,669 individuals, which is approximately half of the no testing peak value without social distancing. This finding is not surprising given that we model social distancing by reducing the contact rate *β* by half. Generally, social distancing expands the range of testing capacities over which the peak can be reduced to its initial value for a given information level. Compare, for example, the *η* = 0.90 curve in Fig. 3c to that of Fig. 1a. We thus conclude that social distancing allows for more effective utilization of limited testing capacities under lower information quality levels. Note that in both the base and socially distanced parameter scenarios, we find no non-zero testing capacities for which the peak can not be suppressed to its initial size for *η* = 1, and therefore no range of testing capacities over which the optimal *ρ* is unique (Figs. 3d and 1b).

Combining the two modulating factors shows that the beneficial effects of social distancing at low testing capacities can counteract some of the detrimental effects of delays in testing implementation (Figs. 3e and 3d). Indeed, social distancing reduces the testing capacity range over which implementation delays render epidemic control impossible. This interval is given by [0, 0.4] tests per thousand per day with 50% contact reduction social distancing (Fig. 3e), as compared to [0, 1.2] without social distancing (Fig. 3a). For all delays between 1 day and the time of the uncontrolled epidemic peak, 62 days, larger degrees of contact reduction from social distancing yield larger reductions in the range of testing capacities for which the peak can not be reduced to its initial size under perfect information (Fig. 4). Note that after day 62, the infection curve turns downward in the uncontrolled model, so for delays greater than 62 days in the controlled model, the epidemic peak value will always be equal to the initial value regardless of testing capacity, and peak reduction is not a useful control goal. In total, these results emphasize the importance of social distancing during the early resource-limited stages of a novel disease epidemic.

**Figure 4:**
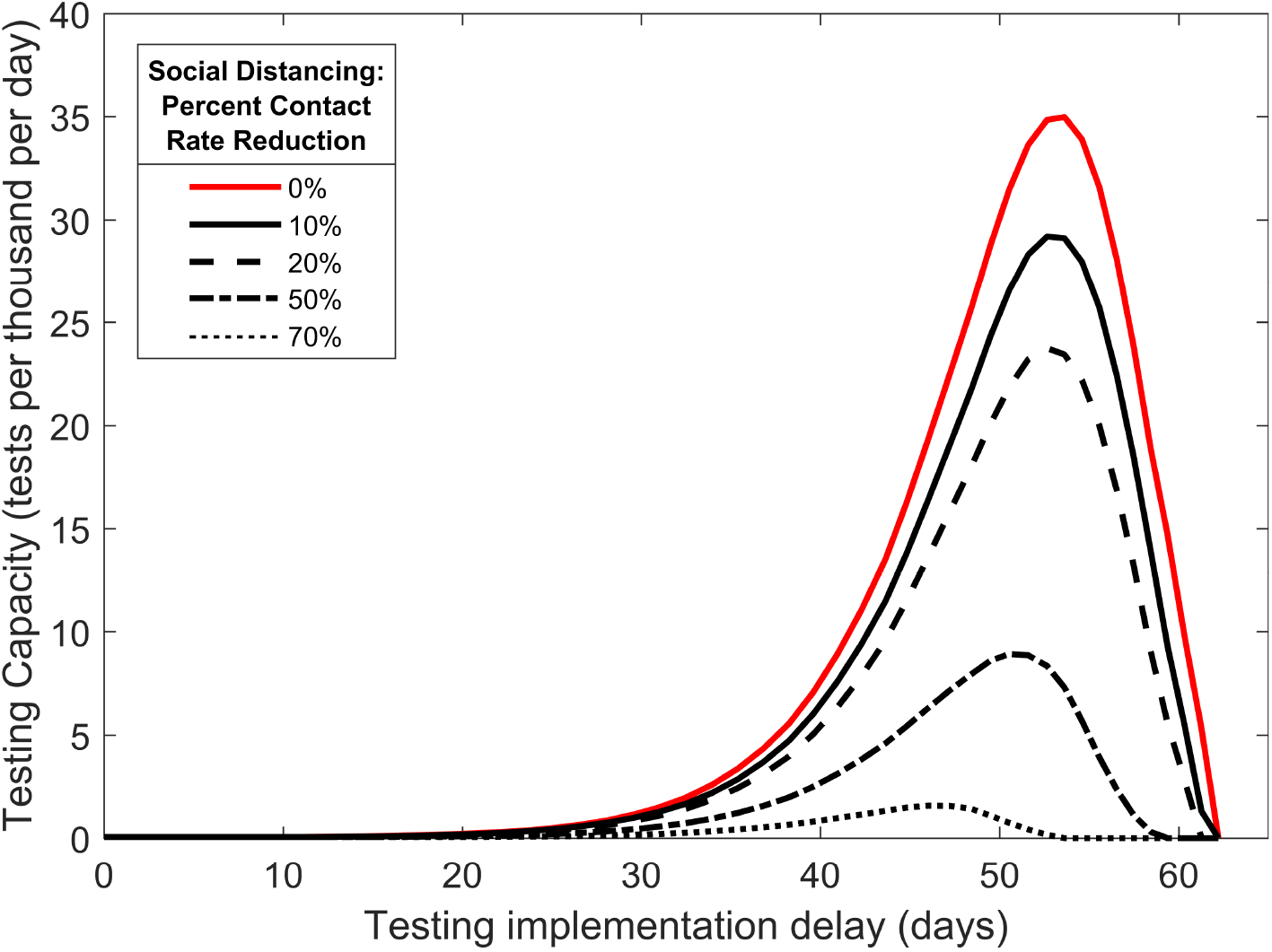
Combined effects of social distancing and delays in testing implementation on epidemic controlability. Threshold testing capacities are plotted as a function of implementation delay, where different curves represent different social distancing strengths as percent reduction in the contact rate. For a given implementation delay time, if testing capacity falls below the value indicated by a curve in the figure, the epidemic will not be forced into a downturn upon control implementation despite perfect information, assuming the indicated level of social distancing.

## Discussion

The COVID-19 pandemic has exposed a critical lack of capacity for diagnostic testing in an emerging pandemic. Using a modified SEIR model, we explored how distributing a limited amount of testing effort can affect the course of an epidemic when testing is directly coupled to quarantine. The model is tailored to the epidemiology of SARS-CoV-2, and divides infected individuals into symptomatic and non-symptomatic classes, with the latter class including individuals who have been exposed but are not yet infectious as well as those who are infectious but not strongly symptomatic. We further defined clinical testing as that focused exclusively on the symptomatic class, while non-clinical testing is distributed across the rest of the potentially infected population (S, E, A, and U). For a given testing capacity *C*, our model thus allows us to identify optimal testing policies in terms of the balance between clinical and non-clinical testing, modulated by the strategy parameter, *ρ*, and the information parameter *η*. This latter parameter governs the extent to which non-clinical testing is concentrated on infected individuals. We further examined how optimal policies shift as a function of testing capacity.

Focusing on the goal of maximally reducing the height of the infection curve (i.e., “flattening the curve”), we found that optimal testing is always able to supress the epidemic, provided that testing is implemented at the onset of disease transmission. Clinical testing strategies are generally optimal at low testing capacities. Under some conditions when the testing rate is low, mixed strategies that include a small but finite amount of non-clincal testing are optimal, but only when there is essentially perfect information with which to focus non-clinical testing on infected individuals. While perfectly omniscient non-clinical testing is unlikely to be achieved in reality, high *η* values may be possible with efficient contact tracing and case investigation programs. These results therefore suggest that testing policies employed in many countries early in the pandemic, which strongly emphasized clinical testing with some additional testing effort aimed at the highest risk individuals (e.g., front-line healthcare workers), were reasonable. Furthermore, we demonstrated that exclusively non-clinical testing is never the optimal strategy. In other words, non-clinical testing plus a small but finite amount of clinical testing will always be better than a purely non-clinical strategy for epidemic peak reduction.

Since the onset of the pandemic, testing capacity has steadily increased throughout much of the world. Our results show that increased testing capacity brings with it a broader range of possibilities for optimizing testing. As testing rate increases, the amount of proxy information required for a mixed strategy to be optimal decreases, with all other factors held constant. At a testing capacity of 8 tests per day per 1000 people, a mixed strategy becomes optimal even when there is no proxy information available to pinpoint non-symptomatic infected individuals. This testing rate thus defines the minimal testing capacity for which a broad, non-targeted population monitoring program, in conjunction with clinical testing, is optimal. While on the higher end of the realistic range of testing rates, this level of testing has been exceeded in several countries, including Denmark, Iceland, Luxembourg, and United Arab Emirates.

While we have chosen minimizing the height of the peak of the infection curve as a pragmatic and meaningful control goal, we also explored the common approach of minimizing *R*_0_ (see Appendix). A mathematical advantage of *R*_0_ minimization is that it leads to closed-form expressions for key threshold parameter values that delimit the conditions under which different testing strategies are optimal. However, we found that for our model, results between these two control goals often differed markedly. Specifically, we identified conditions under which testing policies resulting in *R*_0_ < 1 still yielded large outbreaks, which suggests limited utility of *R*_0_ as a control target. We hypothesize that this phenomenon results from the combination of a finite system size and a finitely small initial condition (see Appendix). We further note that the choice of control goal can also lead to qualitatively different conclusions about optimal strategies. For example, purely clinical testing strategies are never optimal under *R*_0_ minimization, which contrasts sharply with low testing capacity results for peak minimization.

Our results suggest that testing early is critically important to control efforts. Specifically, the range of testing rates that allows full epidemic control is broadest when testing is implemented immediately at the start of an epidemic. A delay of even 30 days is sufficient to significantly narrow the conditions under which the epidemic can be brought to heel. Looking in the other direction, mitigation efforts that lower the effective contact rate, such as lockdowns, social distancing, and mask wearing, significantly facilitate epidemic control, particularly when combined with early testing. Importantly, interventions that reduce the contact rate also lower the threshold testing capacity where uniform random testing of the non-symptomatic population is warranted. These considerations suggest that testing programs should designed in conjunction with non-pharmaceutical interventions.

Taken together, our results suggest that focusing exclusively or mostly on clinical testing at very low testing capacities is often optimal or close to optimal. As testing capacities increase, which can typically be expected to happen with time since epidemic onset, the options for optimally distributing testing effort also open up. To our knowledge, this possibility has been largely unexplored in the literature. This implies that the main gains to be had by optimizing allocation of testing effort will occur at intermediate testing capacities, where options exist for optimization, but capacity is still limited relative to demand. These considerations further suggest that testing policies should evolve over time, and that time-dependent optimal control (Kirk, 1998; Lenhart and Workman, 2007), which can explicitly account for the dynamics of testing capacity, will be necessary to robustly identify how testing should change through the course of an epidemic. While beyond the scope of the present effort, broadening our approach to consider time-depenent optimal control is a clear next step. Another key direction for future efforts would be to consider optimial allocation of testing effort after relaxing the homogeneous, well-mixed population assumption at the core of compartment-type disease models. Spaially explicit extensions of disease models have been shown to change key quantities such as immunization thresholds (Eisinger and Thulke, 2008), and we expect that introducing spatial hetergeneity would also change the picture for optimal testing.

## Data Availability

Only parameter estimates from the literature were used, with values summarized in Table 1, and all sources listed in the literature cited.

## Acknowledgements

This work was partially funded by the Center of Advanced Systems Understanding (CASUS) which is financed by Germany’s Federal Ministry of Education and Research (BMBF) and by the Saxon Ministry for Science, Culture and Tourism (SMWK) with tax funds on the basis of the budget approved by the Saxon State Parliament. This work was also partially funded by the Where2Test project, which is financed by SMWK with tax funds on the basis of the budget approved by the Saxon State Parliament.

## Appendix

In this appendix, we provide an analytic expression for our model’s basic reproduction number, *R*_0_, and we demonstrate that *R*_0_ reduction is not a reliable metric of control efficacy for epidemic peak reduction. The basic reproduction number is a threshold quantity which determines the stability of a disease-free population with no natural or acquired immunity: small numbers of initial cases will produce large epidemic outbreaks when *R*_0_ > 1, and will result in rapid disease die-out when *R*_0_ < 1 (Diekmann et al., 1990). Intuitively, *R*_0_ quantifies the number of secondary cases produced by a typical initial case when interacting with the disease-free state. Because we are able to obtain an analytic expression for *R*_0_, the question of its suitability as a metric for control efficacy is especially prescient; the ability to analytically minimize *R*_0_ rather than numerically minimize the peak itself would provide exact expressions and deep mechanistic insight into optimal control strategies if *R*_0_ were indeed found to be a reliable metric for control efficacy.

### Analytic expression for *R*_0_

The analytic expression for our model’s basic reproduction number is found utilizing the next-generation matrix method (van den Driessche and Watmough, 2002). We find that *R*_0_ can interpreted as the asymptomatic population fraction *f*_*A*_ multiplied by average number of secondary infectious individuals produced by an asymptomatic case, plus the symptomatic population fraction *f*_*Y*_ multiplied by the average number of secondary infectious individuals produced by an symptomatic case:

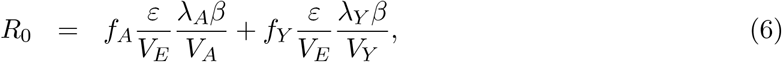

where

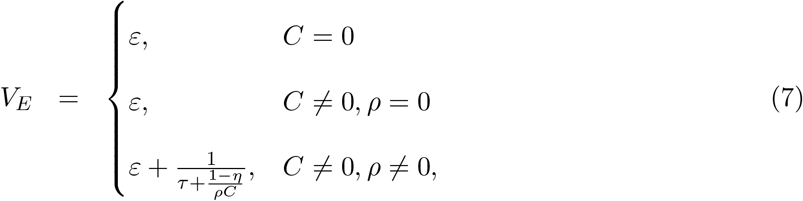

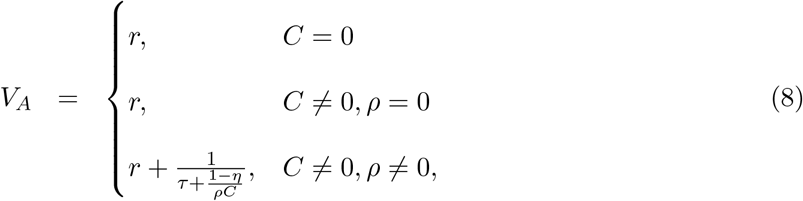

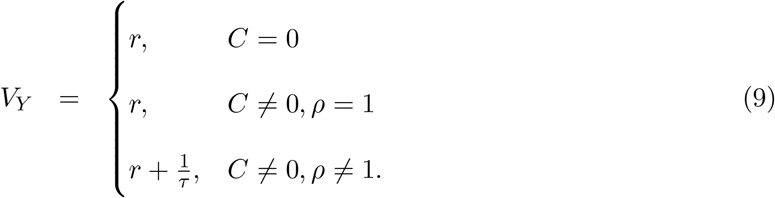

The case *C* = 0 corresponds to the uncontrolled model in Eq. (1), and *R*_0_ is a discontinuous function of *C* at *C* = 0 except for the special case *ρ* = 1, *η* = 1. Under uncontrolled conditions, the parameters in Table 1 give an *R*_0_ = 5.0, with 3.0 originating from the asymptomatic contribution, and 2.0 originating from the symptomatic contribution. For *C* ≠ 0, *R*_0_ is a discontinuous function of *ρ* at *ρ* = 1 and at *ρ* = 0, *η* = 1. Note that these discontinuous limits represent potentially unrealistic extremes of testing policies and information quality.

### Suitability of *R*_0_ as a control metric

To determine if *R*_0_ reduction provides a reliable assessment of control efficacy for epidemic peak reduction, we plot the peak size as a function of *R*_0_ in Fig. 5. These figures were generated by integrating Eq. (5) for specific *C* and *η* and all *ρ* ∈ [0, 1] assuming the baseline parameter values and initial condition, and plotting the resulting peak values against the corresponding *R*_0_ values as determined by Eq. (6). These results show clearly that *R*_0_ is not a reliable measure of control efficacy for epidemic peak reduction, as there exists several cases where the epidemic peak value increases as *R*_0_ decreases. Further, there exist conditions where epidemic peaks are large even though *R*_0_ < 1, in apparent contradiction the definition of *R*_0_ = 1 as a threshold for large epidemic outbreaks. This effect can be seen in for *η* = 1, 0.97, and 0.95 in Fig. 5a, and for *η* = 1 and 0.97 in Fig. 5b. For *η* = 0.97 and 0.99 curves, large peaks occurring with *R*_0_ < 1 correspond to *ρ* values very close but not equal to 1, while for the *η* = 1 curve, correspond *ρ* values very close but not equal to 0.

**Figure 5:**
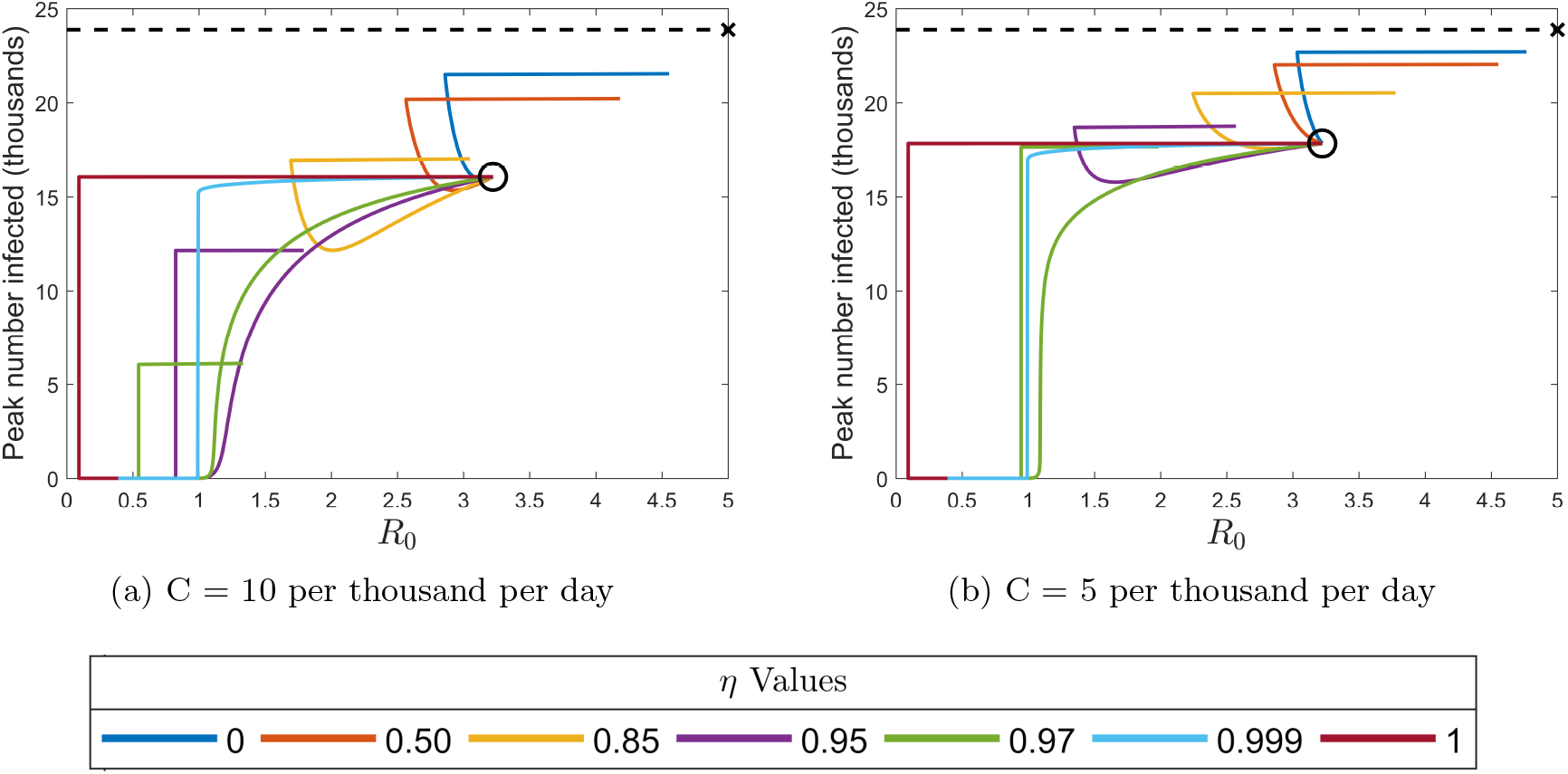
Epidemic peak values plotted as a function of *R*_0_ for testing capacities *C* = 10 and *C* = 5 tests per thousand per day. Curve colors correspond to the information values *η* indicated in the legend. Along each curve, *ρ* increases from 0 and 1, with the beginning of each curve at *ρ* = 0 indicated by the centers of the black circles (*ρ* = 0 represents clinical testing only where the information parameter is irrelevant, so all curves must coincide). The dashed black lines indicate the uncontrolled peak value of 23882 infected individuals, and the black **x** indicates the uncontrolled *R*_0_ = 5.0.

To explain the presence of large outbreaks when *R*_0_ < 1, we define the *effective testing time, τ*_*eff*_, which represents the average time an individual must wait to be tested given the current backlog of patients. For the basic testing model in Eq. (2), the effective testing time is defined by 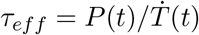, which evaluates to

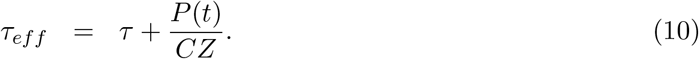

Extending this definition to our disease model with testing and quarantine in Eq. (5), we find two effective testing times for non-clinical and clinical testing, denoted 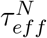 and 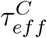, respectively:

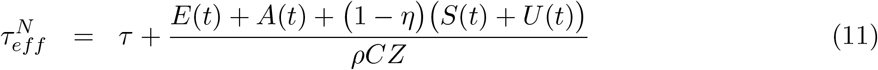

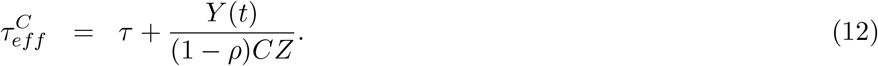

These effective testing times represent the average delays for asymptomatic and symptomatic individuals, respectively, in getting tested, receiving results, and moving to quarantine, given the current backlog of patients and tests. 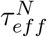 and 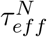 provide measures of non-clinical and clinical control efficiency, respectively, under the current load of infected patients. Specifically, 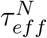 and 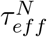 increase monotonically with the patient load (and are thus equal to the minimal possible testing times when the patient load is negligibly small), so for larger patent loads, a fixed number of resources will move individuals to quarantine at a slower effective per-capita rate. In this sense, lower patient loads allow a given number of resources to be leveraged more efficiently.

We hypothesize that the large outbreaks observed when *R*_0_ < 1 arise due to a finite system size and a finitely small initial condition size. The threshold property of *R*_0_ = 1 for outbreak suppression assumes a disease-free equilibrium background state perturbed by a sufficiently small number of initial infected individuals, where “sufficiently small” means small in comparison to the total system size such that the disease dynamics can be well-approximated by linearizing about the disease-free equilibrium. Under disease-free equilibrium conditions, there is no backlog of patients needing to be tested, so the effective testing testing times in Eqs. (11) and (12) achieve their minimal values, and *R*_0_ thus assumes a maximally efficient level of control when assessing outbreak potential. Under the full disease dynamics, however, Eqs. (11) and (12) show that small numbers of initial infectives will produce slightly longer than minimal effective testing times, and that this small increase can become exaggerated when *ρ* is very close but not equal to 1 or 0. Thus, initial conditions can yield testing efficacies much smaller than those assumed by *R*_0_, sometimes to a degree which allows epidemics to grow even when *R*_0_ < 1. In support of our hypothesis, we have found that reducing the initial condition size by a factor of 10 (which corresponds to less than one infected individual) eliminates the effect of large peaks when *R*_0_ < 1 for all cases pictured in Fig. 5.

## Notes

### Competing Interest Statement

The authors have declared no competing interest.

### Clinical Trial

Not Applicable

